# COVID-19–Associated Myocardial Injury and Arrhythmias

**DOI:** 10.1101/2024.11.13.24317287

**Authors:** Mohsin Sheraz Mughal, Hafsa Akbar, Waqar A Mughal, Amit Bansal, Hasan M Mirza, Natale Mazzaferro, Zafar Niaz, Najam Wasty

## Abstract

**Background:** COVID-19 is a systemic illness with broad organ involvement, including the cardiovascular system. While arrhythmias are commonly observed in COVID-19 patients, reports on their incidence, predictors of sudden cardiac death, therapeutic strategies, and both short-term and long-term outcomes remain heterogeneous.

**Methods:** In this retrospective, multicenter study, we evaluated the cohort of patients hospitalized for COVID-19 (RT-PCR confirmed) at Newark Beth Israel Medical Center and Monmouth Medical Center between March 1 and April 30, 2020. This study was approved by the institutional review board. We aimed to assess the incidence of new-onset arrhythmias and major adverse cardiovascular events (MACE), comparing these outcomes in patients admitted to the intensive care unit (ICU) versus those in non-ICU settings.

**Results:** A total of 720 patients were hospitalized for COVID-19 during the study period. The mean age of the cohort was 63.48 years, with 54% male. The incidence of new-onset tachyarrhythmias was significantly higher in the ICU cohort (19%) compared to the non-ICU cohort (8%). The ICU group also experienced a markedly higher incidence of MACE (78% vs. 22%). In-patient mortality was significantly higher in the ICU group (70% vs. 14%). Multivariate regression analysis revealed that ICU admission was an independent risk factor for developing new-onset arrhythmias, with an odds ratio (OR) of 15.92 (95% CI 10.15–24.97; p < 0.001). In patients without MACE, in-hospital mortality was only 1.6%, whereas mortality was 77.4% in those with MACE. ICU admission also emerged as an independent risk factor for MACE, increasing the odds by 15-fold, adjusted for age and gender (OR 15.39, 95% CI 8.36–28.33; p < 0.001).

**Conclusion:** ICU admission is a significant independent risk factor for both new-onset arrhythmias and MACE in COVID-19 patients. The high rates of these adverse outcomes in the ICU population are likely due to the severity of the systemic inflammatory response in COVID-19, rather than a direct viral-mediated cardiovascular effect.

## Background

The novel coronavirus (SARS-CoV-2) infection aka COVID-19 infection is a multi-system disease, affecting nearly all organs and the cardiovascular system is no exception. Studies have suggested that hospitalized COVID-19 patients have a high risk of arrhythmias, and the rate of arrhythmias is even higher among patients admitted to intensive care units. Initial reports from Wuhan, China, noted that up to 16.7% of hospitalized and 44.4% of ICU patients with COVID-19 had arrhythmias. Similarly, myocardial injury is frequently seen in hospitalized COVID-19 patients, with a higher rate of cardiovascular events among patients admitted to intensive care units (ICU). Several mechanisms of COVID-19-related cardiac injury are proposed: oxygen supply-demand mismatch leading to type II myocardial infarction, microvascular injury (dysfunction, or thrombosis), myocyte injury due to direct myocyte invasion, endothelial dysfunction, or indirect inflammation (cytokines) triggered myocyte injury pattern and stress-induced cardiomyopathy aka takotsubocardiomyopathy [2]. Various single-center studies and surveys around the world have documented a spectrum of electrophysiology-related issues associated with COVID-19 such as atrioventricular heart block, atrial fibrillation (AF), polymorphic ventricular tachycardia, and cardiac arrest. Recently, these cardiovascular complications, especially myocardial injury, and arrhythmic manifestations associated with COVID-19 have become more evident [3]. Although arrhythmias appear to be common in COVID-19 patients, there are heterogeneous reports about the rate of arrhythmias. Similarly, arrhythmia mechanisms, characteristics, predisposing factors, their incidence, predictors of sudden cardiac death, therapeutic strategies utilized, as well as short-term and long-term outcomes, and their prognostic implications are not well understood. Hence, we aimed to systematically evaluate the risk of major adverse cardiovascular events (ST-elevation myocardial infarction, non-ST elevation myocardial infarction, sudden cardiac death), and arrhythmias including incidental atrial tachyarrhythmias, ventricular arrhythmias, and bradyarrhythmias. We also interrogated all-cause in-hospital mortality in patients with these cardiovascular manifestations.

## Methods

### Purpose of the Study

Our primary objective was to assess the overall burden of arrhythmias (including incident atrial fibrillation/atrial flutter, bradyarrhythmias, nonsustained ventricular tachycardia (NSVT), and cardiac arrest. We also evaluated the odds of arrhythmias, cardiac arrest, and MACE in patients who required ICU level of care and compared it to the patient population on other medical floors (telemetry).

### Participant selection

Inclusion Criteria: All patients who tested positive for SARS-CoV-2 and were hospitalized during indexed periods were included. A total of 720 patients at both sites (Newark Beth Israel, and Monmouth Medical Center) were included. Exclusion Criteria: Children (age <18 years), and pregnant females were excluded.

### Description of methodology

In this retrospective, multicenter, institutional review board-approved study, we systematically evaluated the urban patient population hospitalized for COVID-19 (RT-PCR confirmed) consecutively admitted at Newark Beth Israel Medical Center, and Monmouth Medical Center from March from 1st to April 30, 2020, in state of New Jersey. Based on requiring ICU-level care patients were divided into two cohorts (ICU vs Non-ICU). ICU admission criteria were hemodynamic instability requiring pressors, or increasing supplemental oxygen requirements (high flow nasal cannula, BiPAP, or requiring mechanical ventilation) In addition, the patient population was divided into two groups based on MACE (ACS: NSTEMI, STEMI, cardiac arrest). We determined clinical outcomes (deceased, still hospitalized, or discharged) at the time of the last follow-up. Demographic characteristics (age, gender, BMI), comorbidities (Supplement, Table-1); the onset of new arrhythmias, and MACE were manually extracted from EMR, and patients with a history of atrial fibrillation were excluded from the new-onset atrial fibrillation analysis. Baseline characteristics were compared between ICU vs Non-ICU patients (Supplement, Table 2). Similarly, baseline features of patients with and without MACE were compared (Supplement, Table 3). Categorical variables are shown in percentages; continuous variables are shown in mean (with standard deviation). Data was extracted manually using the hospital’s electronic medical record. Categorical variables were compared using the chi-square test; continuous variables were compared using the student t-test (paired). *P*-value <0.05 will be considered significant. Baseline characteristics include age, gender, weight, comorbidities, medication history, EKG, and echocardiography. Hematological baseline laboratory findings of CBC, CMP, and troponin I, were collected. In addition, EKG, rhythm strips (scanned in EMR), physicians’ documentation of the cardiac arrest, and echocardiographic findings were also reviewed and collected. A multivariate logistic regression analysis was performed to assess the odds of new-onset arrhythmias, and another model was built to assess the odds of MACE while adjusting for other covariates. Data were collected by the principal and co-investigator(s) and stored on a password-protected computer encrypted to the standards recommended by NBIMC, and MMC IRB. The data stored in the datasheet was de-identified and a unique study ID was assigned. Only this unique study ID and date of the procedure were saved as a key on an encrypted, password-protected computer. The unique study ID was used in the data collection file with no PHI information. If after study data-collection, there was a need to identify individual patients, it could only be done using the unique study ID from the dataset and the separate Master Key. The Master Key was only available to the PI. The Master Key was destroyed after the study completion.

### Statistical Analysis

Proposed sample size: 1,000. We believe about 1,000 patients have been treated at NBIMC and Monmouth Medical Center during the past few months. Approximately 600 patients have been treated at NBIMC. De-identified data will be analyzed using Revman, STATA statistical software, and SAS version 9.4. Continuous variables will be presented as means ± SD, or median (IQR), and range based on normal vs skewed distribution. Shapiro-Wilk tests were performed along with a visual Q-Q plot review to determine the normality of continuous variables. Categorical data will be presented as counts frequencies and percentages. Paired t-test and Wilcoxon rank sum test, Chi-square test, or Fisher’s Exact test to compare variables within patients for continuous and categorical variables when applicable.

## Results

- **ICU vs Non-ICU cohorts**

There were a total of 720 patients hospitalized with COVID-19 during the study period, hospitalized cohort had a mean age of 63.48 years, of these, 54% were male. Only 28% of patients required ICU level of care, while the majority of the study population (72%) were managed on non-ICU medical floors (cardiac telemetry). Patients in the ICU cohort were more likely to be male, obese, and had a history of diabetes mellitus (DM). Baseline characteristics and comorbidities are mentioned in Supplement, Table 2. Patients in the ICU cohort were more likely to have higher levels of troponin I, and HBA1c. New onset tachyarrhythmias were higher in the ICU cohort (19%) vs the non-ICU cohort (8%). New-onset atrial fibrillation was the most common arrhythmia noted, its proportion was nearly two folds higher in the ICU cohort (13% vs 7%). ICU cohort had nearly four times higher proportion of major adverse cardiovascular events (78% vs 22%). All-cause in-patient mortality was profoundly higher in the ICU cohort (70 vs 14%). There were a total of 171 (23.7%) cardiac arrests, of these, only 6% of cardiac arrests were reported on the telemetry floor. The most common rhythm was asystole (12%) followed by PEA (10%); a shockable rhythm was only found in 1.4% of patients (Supplement, Table 2). Multivariate regression analysis showed that the ICU admission factor is an independent risk factor for developing new-onset arrhythmias that increased the odds of new-onset arrhythmias by 15-fold (OR 15.92, 95% CI 10.15-24.97; *p* <0.001) Table 4.

- **Patients with MACE vs Patients without MACE**

A total of 270 patients (37%) developed MACE, regardless of ICU admission status. There were more likely to be older, males with a history of coronary artery disease, heart failure, cardiomyopathy, dyslipidemia, diabetes mellitus, and reported a history of percutaneous coronary intervention. The baseline characteristics and history of comorbidities of both cohorts are mentioned in Supplement, Table 3. Troponin elevation was seen among two-thirds of the patients with MACE, with mean troponin I (max) 4.44 ng/ml (SD 21.1). There were a total of 171 (23.7%) cardiac arrests, of these, the most common rhythm was asystole (12%) followed by PEA (10%); a shockable rhythm was only found in 1.4% of patients. There were 8 cases (3%) of myopericarditis documented. Interestingly, there were 1% strokes, 2.9% pulmonary embolism, and 2.4% deep venous thrombosis noted in this hospitalized cohort. All-cause in-patient mortality in the cohort without MACE was only 1.6%, while in patients with MACE, all-cause in-patient mortality was 77.4%. Multivariate regression analysis showed that the ICU admission factor is an independent risk factor for developing MACE that increased the odds of MACE by 15-fold (Table 5) while adjusted for age, and gender (OR 15.39, 95% CI 8.36-28.33; *p* <0.001).

## Discussion

Invariably, arrhythmias and myocardial injury are frequent manifestations of COVID-19. Data regarding their characterization, predictors, their prognostic implications in hospitalized patients, and the correlation of these cardiovascular events with ICU level of care are equivocal and underpowered. This multicenter study of 720 hospitalized COVID-19 patients in two academic hospitals elaborate COVID-19 associated arrhythmias and myocardial injury–this data revealed important findings:

1. New onset tachyarrhythmias were higher in the ICU cohort (19%) vs the non-ICU cohort (8%). New-onset atrial fibrillation was the most common arrhythmia noted, its proportion was nearly two folds higher in the ICU cohort (13% vs 7%). Logistic regression showed that hat the ICU admission factor is an independent risk factor for developing new-onset arrhythmias that increased the odds of new-onset arrhythmias by 15-fold (OR 15.92, 95% CI 10.15-24.97; *p* <0.001).
2. ICU cohort had nearly four times higher proportion of major adverse cardiovascular events (78% vs 22%). All-cause in-patient mortality was profoundly higher in the ICU cohort (69 vs 14%). Multivariate regression analysis showed that the ICU admission factor is an independent risk factor for developing MACE that increased the odds of MACE by 15-fold while adjusted for age, and gender (OR 15.39, 95% CI 8.36-28.33; *p* <0.001).
3. This study adds that COVID-19 patients with MACE were associated with significant morbidity and mortality, only one-fourth survived to hospital discharge.

Association of ICU with poor outcomes (higher level of arrhythmias, MACE, the total length of stay, and overall all-cause mortality) suggests that these outcomes are likely driven by the severity of illness, a severe systemic inflammatory state; rather than solely by COVID-19 infection [4]. The putative underlying pathophysiological mechanism could be inflammation-driven tissue injury (acute kidney injury) leading to electrolyte derangements [5]. These findings are important to identify the ICU patients with MACE and new-onset arrhythmias, this subset of COVID-19 patients represents poor prognosis as indicated by their higher all-cause in-patient mortality [6]. Several studies have suggested that atrial fibrillation in COVID-19 is associated with unfavorable outcomes, similarly, Identifying these high-risk patients earlier in their course of hospital admission can help healthcare providers to risk stratify them and may opt for aggressive treatment modalities knowing their poor prognosis [7]. At the same time, this data can help clinicians to navigate end-of-life care discussions with the families of this very sick cohort. Compared to other studies this report presents a higher proportion of patients requiring ICU level of care, and higher in-patient all-cause mortality suggesting that our study population represents a sicker population [6, 8, 9]. This study has several limitations; retrospective study design, lack of ethnic, and socioeconomic information, missing laboratory findings, and lack of continuous telemetry data led to an inability to assess subclinically arrhythmias. In addition, our analysis was limited to only hospital-course, we lacked a complete out-patient follow-up. There is inconsistency in the definition of MACE across the board, some studies include strokes and heart failure events in MACE. This data represents a larger urban population at two academic hospitals in New Jersey during the initial surges of COVID-19, hence, these findings may not be generalizable to the current vaccinated population hospitalized with COVID-19.

## Conclusion

ICU admission is an independent marker of MACE and new onset arrhythmia in COVID-19; it independently increased the odds of new-onset arrhythmias and MACE by 15-fold. Based on this data, the high rate of MACE, and new onset arrhythmias in the ICU population is likely a consequence of severe systemic inflammatory illness rather than a direct viral infection-related cardiovascular manifestation.

## Clinical implication

Our data suggest that ICU admission in patients with COVID-19 is associated with a significantly increased risk of major adverse cardiovascular events (MACE) and new-onset arrhythmias. Specifically, ICU admission independently increases the odds of experiencing new-onset arrhythmias and MACE by 15-fold.

The clinical significance of this information includes the following:

1. Identification of High-Risk Patients: ICU admission serves as a marker for identifying COVID-19 patients at high risk for cardiovascular complications. Clinicians should be vigilant in monitoring these patients for signs and symptoms of MACE and arrhythmias.
2. Risk Stratification: The data underscores the importance of risk stratification in COVID-19 patients, particularly those admitted to the ICU; risk stratification allows for targeted interventions and closer monitoring of high-risk patients.
3. Understanding Disease Pathophysiology: The findings suggest that the high rate of MACE and new-onset arrhythmias observed in ICU patients with COVID-19 may be attributed more to severe systemic inflammatory illness rather than direct viral infection-related cardiovascular manifestations. This insight into disease pathophysiology can inform treatment strategies and guide research efforts aimed at mitigating cardiovascular complications in severe COVID-19 cases.
4. Clinical Management: Clinicians caring for ICU patients with COVID-19 should be prepared to manage cardiovascular complications effectively. This may involve early recognition and treatment of arrhythmias, aggressive management of underlying systemic inflammation, and coordination with multidisciplinary teams to optimize patient outcomes.

Overall, the data highlights the significant cardiovascular implications of severe COVID-19 illness and underscores the importance of comprehensive cardiovascular assessment and management in ICU patients with COVID-19.

## CRediT authorship contribution statement

Mohsin Sheraz Mughal: Conceptualization, Methodology, Data curation, Writing - original draft, Writing - review & editing, Formal analysis, Analysis, Writing-review & share first authorship, had full access to all the data in the study and take responsibility for the integrity of the data. Amit Bansal: Conceptualization, Methodology, Data curation, Writing - original draft, Analysis, Writing-review & share the first authorship, had full access to all the data in the study and take responsibility for the integrity of the data. Hasan Mirza: Writing - review & editing. Zafar Niaz: Writing, and literature review. Statistical analysis: Natale Mazzaferro. Afzal Ur Rehman: Review & editing. Najam Wasty: Review & editing. Mohsin S Mughal: Project administration, Supervision, Validation, had full access to all the data in the study and took responsibility for the integrity of the data. Waqar A Mughal, writing draft, and literature review.

## Data Availability

All data produced in the present study are available upon reasonable request to the authors

## Acknowledgment

We thank Saba Ahmed, MD; Priyanka Singh, MD; Waqar A. Mughal, M.B.B.S.; and Zahid Ali Raza, M.B.B.S. for their assistance in the literature review.

## Notes

### Competing Interest Statement

The authors have declared no competing interest.

### Funding Statement

None

### Author Declarations

IRB committee Rutgers Health Newark Beth Israel Medical Center; Rutgers Health Monmouth Medical Center

